# Is socioeconomic status associated with risk of childhood type 1 diabetes? A comprehensive review

**DOI:** 10.1101/2022.06.21.22276686

**Authors:** Paz Lopez-Doriga Ruiz, Lars C. Stene

## Abstract

**Aims:** Studies of social inequality in risk of type 1 diabetes seems inconsistent. The present review aimed to comprehensively review relevant literature and describe what has been reported on socioeconomic status or parental occupation and risk of type 1 diabetes in children.

**Methods:** We searched for publications between January 1, 1970, and November 30, 2021. We focused on the most recent and/or informative publication in case of multiple publications from the same data source and referred to these as primary studies.

**Results:** Our search identified 69 publications with relevant data. We identified eight primary cohort studies with individual-level data, which we considered the highest quality of evidence. Furthermore, we identified 13 primary case-control studies and 14 semi-ecological studies with area-level socioeconomic status variables which provide weaker quality of evidence. Four of eight primary cohort studies contained data on maternal education, showing non-linear associations with type 1 diabetes that were not consistent across studies. There were no consistent patterns on the association of parental occupation and childhood-onset type 1 diabetes.

**Conclusions:** There is a need for more high-quality studies, but the existing literature does not suggest a major and consistent role of socioeconomic status in the risk of type 1 diabetes.

**Novelty statement:** *“What is already known?”:*

- Socioeconomic status has been associated with a variety of exposures, but the influence on type 1 diabetes risk is unclear.

*“What this study has found?”:*

- Our searches identified eight high-quality and several lower quality studies, mostly using socioeconomic status as a confounder. There was no consistent association between socioeconomic status and risk of childhood type 1 diabetes. No conclusions could be drawn for specific parental occupations.

*“What are the implications of the study?”:*

- While there is a need for more high-quality studies, the existing literature does not suggest a major and consistent role of socioeconomic status in the risk of type 1 diabetes.

## Introduction

The incidence of childhood-onset type 1 diabetes varies widely between countries and tend to be more common in wealthier countries (1). The incidence has doubled during two to three decades in many countries (2). Environmental factors, probably operating in early life, are therefore likely involved in the aetiology (3).

Lower parental socioeconomic status has been consistently associated with a variety of lifestyles and exposures hypothesized to be linked to the risk of childhood-onset type 1 diabetes such as maternal and child obesity, smoking in pregnancy, lack of breastfeeding, child infections (4-6). Studies describing risk of type 1 diabetes according to socioeconomic status can therefore shed light on the aetiology of type 1 diabetes (Figure 1).

**Figure 1.**
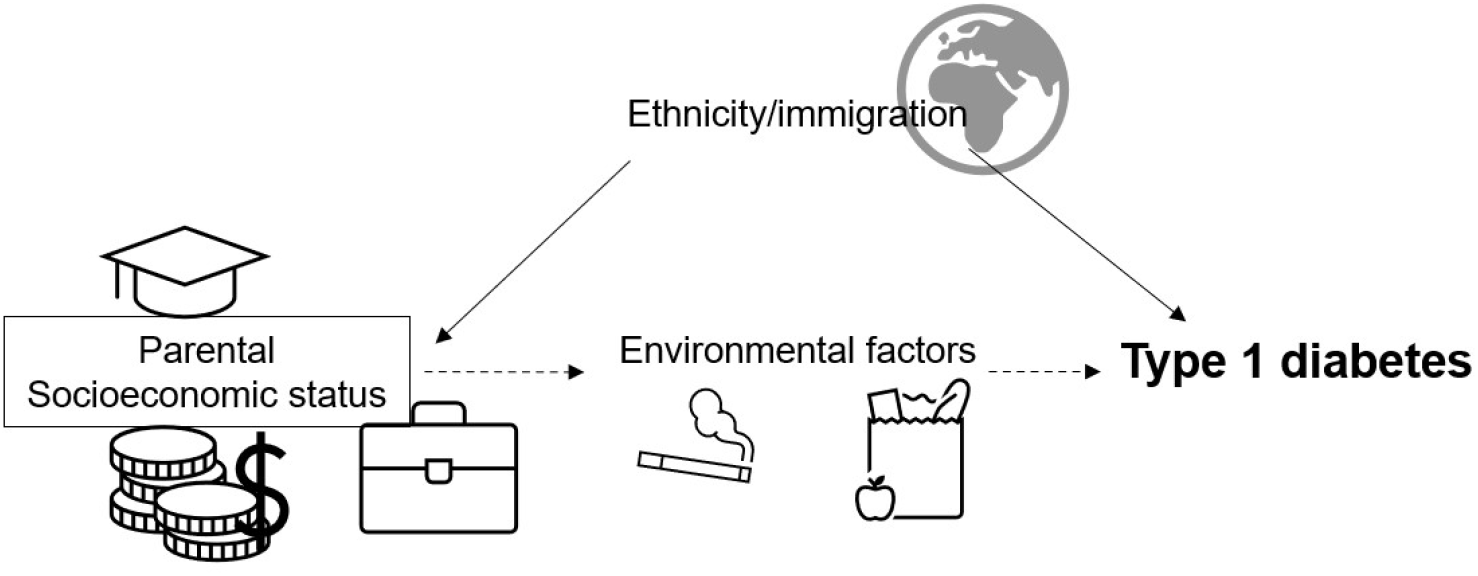
Illustration of the research question. Potential role of socioeconomic status as a risk factor of type 1 diabetes.

Socioeconomic status variables may have different meanings and interpretations in different locations and at different times. For health outcomes in children, it is the parental socioeconomic status that is relevant. Education, occupation, and income has traditionally been the most frequently used measures in epidemiology (7-9). Occupation has traditionally been used in some countries to categorise families in groups of social status. Specific parental occupations may serve as indicators of exposures, including prenatal exposures, that may provide clues to the aetiology of type 1 diabetes. For instance, studies in asthma and allergy have suggested that farm living is associated with lower risk of these outcomes (10). Teachers or health workers are typically frequently exposed to infections (11, 12). Industrial workers may be exposed to toxic chemicals (13).

The literature linking socioeconomic status or parental occupation and risk of type 1 diabetes is scattered and is rarely mentioned in reviews of risk factors for the disease. A 1982 review suggested higher risk of type 1 diabetes in children from families with high socioeconomic status (14). In contrast, a 2010 review of type 1 diabetes epidemiology emphasized that there were inconsistent methods and results across studies (15), and a 2014 review of socioeconomic status and autoimmune disease briefly covered type 1 diabetes (16). An updated review of this topic is lacking.

We therefore aimed to comprehensively review relevant literature on the relationship between socioeconomic status or parental occupation and the risk of childhood onset type 1 diabetes.

## Methods

### Search strategy and inclusion criteria

We carried out a comprehensive literature review of socioeconomic status and the potential association with incidence of childhood-onset type 1 diabetes. We searched PubMed for publications between January 1, 1970, and November 30, 2021 (search terms in Supporting Information). We focused on the most recent and/or informative publication in case of multiple publications from the same data source and referred to these as primary studies. We excluded all studies of socioeconomic consequences of type 1 diabetes. Including prevalent cases of type 1 diabetes was considered a methodological weakness, especially if socioeconomic status variables were only available after diagnosis, because having a child with type 1 diabetes may influence parental socioeconomic status.

We imported the identified articles to the software Covidence© and the duplicates were characterized and excluded. We also included articles from our personal reference lists from a previous review (17) and references from review-articles from 2010 and 2014 (15, 16). In addition, we assessed studies included in previous systematic reviews on risk factors of type 1 diabetes (18, 19). We included studies with data on socioeconomic variables and incident type 1 diabetes during childhood (age <18 years) and focused on the most recent and/or informative publication in case of multiple publications from the same data source. The reference lists from publications with a main aim of investigating socioeconomic status in relation to risk of type 1 diabetes were screened for additional publications. Studies with a minimum of 100 cases of incident type 1 diabetes were included. Both authors screened the articles, and we resolved any disagreement through discussion.

### Study designs and quality of evidence

Study designs were categorized based on whether individual level childhood socioeconomic status or area-based socioeconomic status was available, and on whether the study design was cohort, case-control, ecological, or other. Cohort studies with detailed individual level information on were considered the highest-level evidence, particularly if based on complete population-based registries. Case-control studies nested within registries, without need for active participation in an interview or returning a questionnaire, were considered equal level evidence to that for a cohort design. Traditional case-control studies have a number of potential limitations, and even more so for ecological studies (see discussion section). Ecological study designs were considered the lowest quality of evidence. A study was labelled ecological if socioeconomic status was only available at level of area of residence (even if type 1 diabetes were available at the individual level, see Supporting Information for details).

## Results

After screening 240 titles/abstracts from the PubMed search and excluding the majority due to lack of relevant data, 35 publications with relevant data were assessed in detail. We added 34 additional publications from other sources to a total of 69 publications with relevant data (Figure 2).

**Figure 2.**
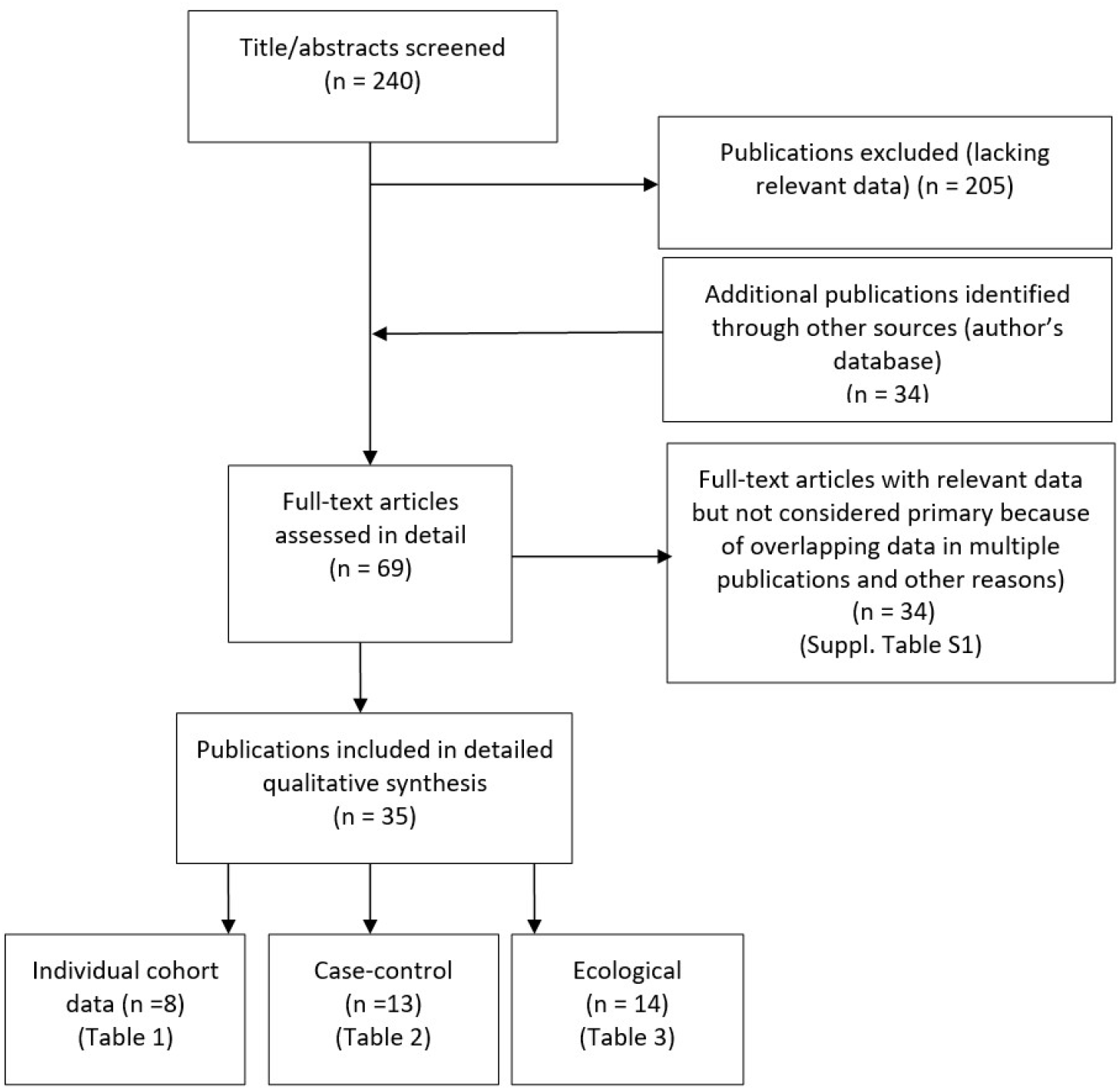
Flow chart of literature review.

Eight primary publications used cohort design (including one large scale registry-based case-control study considered of equivalent quality as cohort) with individual level data on socioeconomic status (Table 1). Thirteen primary case-control studies are presented in Table 2. All case-control studies had individual level socioeconomic status data (two had area-based socioeconomic status in addition). Fourteen primary ecological studies are presented in Table 3.

**Table 1.** Literature review: Primary cohort studies of individual level socioeconomic status and incidence of type 1 diabetes*

| First author, year published (reference) | Period T1D diagnosed | No. w/T1D (age)† | Place | SES variables‡ | Direction of association§ | Results§ | Comments |
| --- | --- | --- | --- | --- | --- | --- | --- |
| Begum 2020 (32) | 2002-2014 | 333 (<11 y) | South Australia | Parental employment status, public/private hospital (+ area-based) | + | Both parents employed vs none: uRR=1.3 (unclear statistical significance) | Only unadjusted associations shown |
| Bengtsson 2020 (21) | 1980-2015 | 8335 (<35 y) | Denmark | Family poverty (5.5% of cohort), parental long-term unemployment (25% of cohort) | No clear association | RRs 0.94-1.05, not significant | SES part of “adversities” before age 18, as time-varying |
| Syrjälä 2019 (33) | 1996-2018 | 188 | Finland (Oulu and Tampere) | Maternal education | Non-linear | uRR for highest to lowest mat educ., 1.00, 0.63 (upper vocational), 0.55 (vocational) | Cohort with genetically susceptible children |
| Clausen 2016 (34) | 1997-2012 | 1503 | Denmark | Mat. and pat. educ. at child's birth (incl vocational/occupation) | Non-linear | Highest risk in mid of 3 categories for mat.educ (uRR=1.13, 95%CI 0.99-1.29), no assoc. with paternal education | Ordering of levels not obvious\|\| |
| Khashan 2015 (35) | 1973-2009 | 13944 | Sweden | Maternal education | Non-linear | Highest risk in mid cat vs low: uRR=1.1; high vs low: uRR=0.9 (both significant) | uRR calculated from Table 1 |
| Bruno 2013 (22) | 1984-2007 | 274 | Italy (city of Turin) | Parental education, crowding | + (non-linear) | Age 4-14y: high vs low par. educ.: RR=1.67 (mid vs low: uRR=0.99). Low vs. high h.hold crowding: RR= 0.83 (95%CI: 0.56-1.22) (educ. and crowding mutually adjusted) | Other ages studied but no significant parental educ. x age interaction |
| D'Angeli 2010 (36) | 1992-2005 | 899 (<19 y) | US (Washington State) | Maternal education | + (non-linear) | Mat. educ. high (>=13y) vs low (<12y): aRR= 1.9 (p<0.05). Mid (12y) vs low: aRR=1.8. | Assoc. adj. for maternal age and marital status. Mat educ. only for births after 1992 |
| levins 2007 (37) | 1963-1999 | 348 | UK (Oxfordshire, West Berkshire) | Maternal social class | No clear association | High (I and II) vs. low (IV and V): uRR=1.2, not significant | 20% missing data |

u/aRR: Unadjusted (u) or adjusted (a) relative risk (or odds ratio, incidence rate ratio or hazard ratio) comparing higher versus lower socioeconomic status. H.hold: Household.
\* One register-based, large scale nested case-control study was considered of equal quality as cohort design (D'Angeli 2010). Bruno 2013 was the only publication where investigation of socioeconomic status (SES) as a risk factor for type 1 diabetes (T1D) was the main aim, while socioeconomic status was only a covariate in Syrjälä 2019, Khashan 2015, and levins 2007. The following had accounted for ethnicity by adjustment or restriction: Bengtsson 2020, D'Angeli 2020 and possibly Begum 2020.
† Age (years) at diagnosis of type 1 diabetes (T1D) less than 15 years unless otherwise specified. Cases with available data on socioeconomic status are reported.
‡ Socioeconomic (SES) variables. Parental educ (education) or occupation refers to highest level within the pair of maternal or paternal education, as opposed to separate data for mat. (maternal) or pat. (paternal) educ.
§ Comparing higher versus lower socioeconomic status (SES). +: increased socioeconomic status was associated with higher risk of type 1 diabetes. -: Increased socioeconomic status was associated with lower risk of type 1 diabetes. For socioeconomic status variables such as deprivation and unemployment mean low socioeconomic status. Socioeconomic status based on occupation typically used in the UK are often labelled with roman number I for high social class and III, IV or V refers to lower socioeconomic status.
|| The three levels of maternal and paternal education in Clausen 2016 were labelled "Elementary school/high school", "Short education/skilled worker", and "Medium/long education". It is not obvious whether skilled worker refers to education or actual occupation, and the ordering of these categories is not entirely obvious.

**Table 2.** Characteristics of 13 primary case-control studies of socioeconomic status and T1D*.

| First author, year published (reference) | Period T1D diagnosed | No. w/T1D (age)† | Place | SES variables‡ | Direction of association§ | Results | Comments |
| --- | --- | --- | --- | --- | --- | --- | --- |
| Liese 2012 (38) | 2003-2006 | 507 (10-22y) | US (2 centres) | Parental educ. and income, area characteristics | + | Parental high vs low educ: uRR=2.1; high vs low income uRR=4.9 | Response in eligible controls: ca 20% |
| Karavanaki 2008 (39) | 1999-2000 | 107 (<16y) | Greece (Athens) | Mat. and pat. educ. and occup | – | Mat. university vs elem. uRR=0.1; Pat. univ vs elem uRR=0.51, n.s.) Occup socioeconomic status not consistent (n.s.) | Also reported socioeconomic status but not defined. Small study |
| Svensson 2005 (40) | 1996-1999 | 490 | Denmark | Maternal white collar worker or not | – | Mat. white collar worker (high socioeconomic status) vs not (low): aRR=0.82 (95%CI: 0.64-1.05) | Response: 81% in cases and ca 48% om controls |
| Sipetic 2004 (41) | 1994-1997 | 105 (<16y) | Serbia (Belgrade) | Mat. and pat. educ. and occupation | + (non-linear) | Mat. educ high vs. low: uRR=1.1 (n.s.) Pat. educ. high vs. low uRR=1.6 (sign.). Highest T1D risk in mid cat. | Controls hospitalised. Small study. Pat educ. sign. after adjustment in Sipetic 2005 (42) |
| Stene 2003 (23) | 1997-2000 | 545 | Norway | Mat. educ. | (–) | Mat.educ highest vs lowest: aRR=0.60 (CI: 0.32-1.10) | Response: cases 73%, controls 56%. |
| Wadsworth 1997 (43) | 1992 | 218 (<5y) | UK and Ireland | Mat. and pat. soc. class | No clear assoc. | Mat. class high vs low: uRR=0.89 (n.s.); pat class: uRR=0.80 (n.s.) | Only <5-year-old children |
| McKinney 1997 (44) | 1993-1994 | 196 | UK (Yorkshire) | Mat. educ. and occup. soc. status | No clear assoc. | Mat educ vs none: high: RR=0.84, basic: RR=0.79 (n.s.). No assoc with mat occup |  |
| Verge 1994 (45) | 1990-1991 | 235 | Australia (New South Wales) | Mat. educ. | – | Mat. educ high vs none: uRR ca 0.6, p(trend)=0.06 | Response 92% in cases, 55% in controls |
| Soltész 1994 (46) | 1990 | 130 | Hungary | Mat. educ. | – | Mat educ university vs not: uOR=0.59 (p=0.07) | Friends as controls matched for socioeconomic status to some extent |
| Patterson 1994 (47) | 1975-1988 | 529 | N. Ireland and Scotland | Pat. occup. (soc. class) | Inconsistent | High pat. occup. soc. class->higher T1D in N. Ireland, but n.s. lower T1D in Scotland. |  |
| Lawler-Heavner 1994 (48) | 1978-1988 | 221 | US (Colorado) | Mat. educ., fam. income | – | Mat >high school vs high school or less: uRR=0.63 (p=0.03). fam.income high vs low: uRR=0.49 (p[trend]<0.001) | Results seems inconsistent with overlapping data in Mayer 1988 (Table S10). |
| Virtanen 1991 (49) | 1988-1989 | 103 (<7y) | Finland | Mat. educ. | – | >13 y of educ vs ≤13y: uRR=0.51, p=0.04 | Response 95% in cases, 54% in controls. Small study. |
| Blom 1989 (50) | 1985-1986 | 339 | Sweden | Mat. and pat. educ. and occup. | – | Mat university vs elementary: uRR = 0.66 (p<0.05) (pat educ. similar but n.s.). Pat self- employed vs manual: uRR ca 0.7-0.8 | Response 86% in cases, 67% in controls |

u/aRR: Unadjusted (u) or adjusted (a) relative risk (or odds ratio, incidence rate ratio or hazard ratio) comparing higher versus lower socioeconomic status (SES). Often calculated from characteristics tables in publications. socioeconomic status: Socioeconomic status.
\* Case-control studies where controls (and typically also cases) were invited to participate for data collection via questionnaire (ten studies) or interview (two studies: McKinney 1997 and Liese 2012). Patterson 1994 used information from health records. All studies typically have less than complete participation, and lower participation among controls than among cases was quite common.
† Age (years) at diagnosis of type 1 diabetes (T1D) less than 15 years unless otherwise specified. Cases with available data on socioeconomic status are reported.
‡ Socioeconomic (SES) variables. Parental educ (education) or occupation refers to highest level within the pair of maternal or paternal education, as opposed to separate data for mat. (maternal) or pat. (paternal) educ.
§ +: increased socioeconomic status was associated with higher risk of type 1 diabetes. –: Increased socioeconomic status was associated with lower risk of type 1 diabetes. Lower level of deprivation and unemployment means higher socioeconomic status. Socioeconomic status based on occupation typically used in the UK are often labelled with roman number I for high social class and III, IV or V refers to lower socioeconomic status.

**Table 3.**
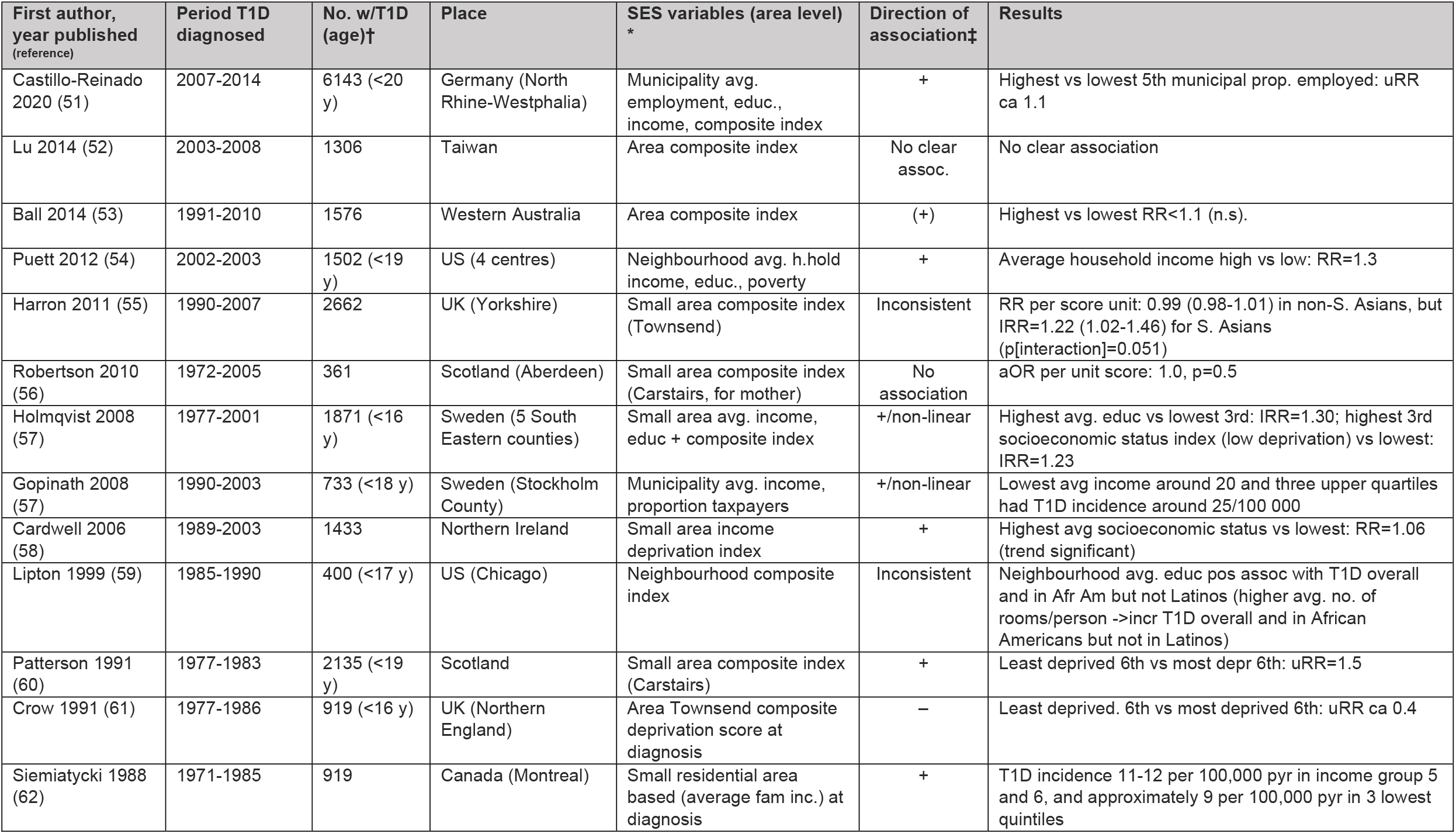
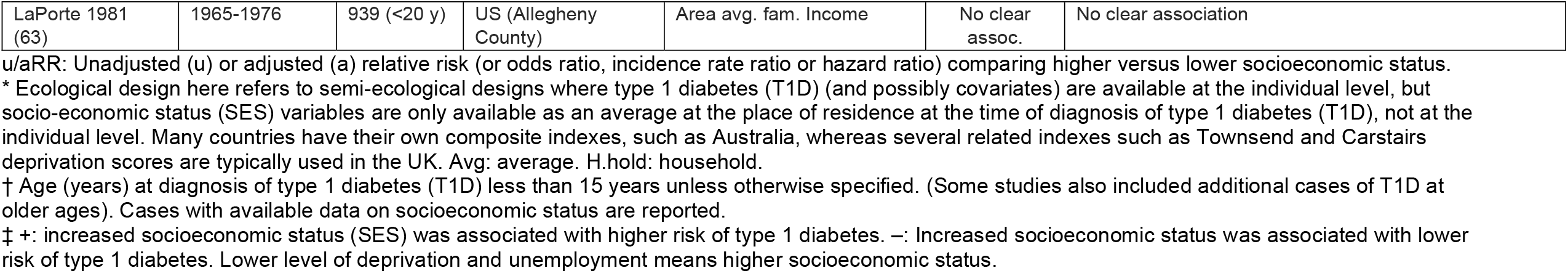
Characteristics of 14 primary ecological studies of socioeconomic status and T1D*.

The majority of studies had not accounted for ethnicity or country background, which may lead to confounding. The majority of the cohort and case-control studies included socioeconomic status as an adjustment variable, not as a primary study variable. A summary of the numbers of publications with different strengths and weaknesses are presented in the Supporting Information results section. A meta-analysis was not possible due to heterogeneity of the socioeconomic indicators, but major studies were tabulated and characterized in terms of main characteristics and direction of association (20).

### Maternal or paternal education in relation to risk of type 1 diabetes

Four of eight primary cohort studies contained data on maternal education in relation to risk of type 1 diabetes, showing non-linear associations with highest risk of childhood-onset type 1 diabetes in the mid (or highest) of three categories of maternal education in one, and a U-shaped association in one study (Table 1).

Of 14 primary case-control studies, nine contained data on maternal education, two on paternal education and one on parental education (highest of maternal or paternal education). Of the nine studies with maternal education, six reported inverse association, and the remaining showed no clear association. Of two studies with paternal education, one showed a positive and the other no significant association with type 1 diabetes (Table 2).

### Occupation-derived socioeconomic status and risk of childhood type 1 diabetes

One primary cohort study reported maternal social class based on occupation and found no significant association. Parental unemployment tended to be associated with higher risk of type 1 diabetes in a study from Southern Australia, but not in a study from Denmark (Table 1).

Of 14 primary case-control studies, six reported social class according to maternal occupation, and six according to paternal occupation, and there was no clear association with childhood-onset type 1 diabetes in these (Table 2).

### Parental income and risk of childhood type 1 diabetes

None of the primary cohort studies reported associations for parental income (main Table 2). Parental employment status (both parents working, only father, only mother or neither), use of public versus private hospital were reported by Begum et al. in South Australia, showing a slightly but significantly higher risk associated with higher socioeconomic status. In a Danish study, family poverty (5.5% of cohort), parental long-term unemployment (25% of cohort) was not significantly associated with childhood-onset type 1 diabetes (21).

Two of the 14 primary case-control studies contained data on parental or family income, both from the USA, and results showed associations in the opposite direction (Table 2).

### Other socioeconomic status indicators and risk of type 1 diabetes: Crowding

Bruno 2013 analysed household crowding (persons per area of the residence) in the city of Turin, Italy, and found no significant association in the 4–14-year-olds (Table 1). Bruno found higher crowding index to be associated with a borderline significantly higher risk of type 1 diabetes in the 0–3-year-olds (22).

### Area-based socioeconomic status in relation to type 1 diabetes incidence

Details regarding methodological aspects and composite indices used in area-based studies are described in Supporting Information results section. Five of 14 primary studies analysing area-based based socioeconomic status in relation to type 1 diabetes incidence found a positive relation, while one found a clear inverse relation (Patterson 1991) and the remaining found no clearly significant associations or suggestive non-linear associations (Table 3).

### Occupation-derived socioeconomic status and risk of childhood type 1 diabetes

One primary cohort study reported maternal social class based on occupation and found no significant association. Parental unemployment tended to be associated with higher risk of type 1 diabetes in a study from Southern Australia, but not in a study from Denmark (Table 1).

Of 14 primary case-control studies, six reported social class according to maternal occupation, and six according to paternal occupation, and there was no clear association with childhood-onset type 1 diabetes in these (Table 2).

### Specific maternal or paternal occupation in relation to type 1 diabetes

We found no study investigating risk of childhood-onset type 1 diabetes across several types of maternal or paternal occupations. However, we found three studies of some relevance for parental farming, showing no clear association with risk of childhood type 1 diabetes (see Supporting Information results section for details).

We found only publication investigating parental occupational exposure, a case-control study from which it was difficult to draw firm conclusions (see Supporting Information results section for details).

## Discussion

There were remarkably few high-quality studies relating socioeconomic status or parental occupation to childhood type 1 diabetes. Many studies reported non-linear associations, but there was little or no consistency across studies, even among the highest quality studies.

### Methodological weaknesses in published studies

Several methodological weaknesses were apparent in the majority of studies assessed. Registry-based case-control studies typically do not require consent or active participation, or at least consent and/or participation in data collection is done before the disease outcome and hence similarly for cases and controls. On the other hand, traditional case-control studies require active participation and usually involves collection of data at or after diagnosis of cases with type 1 diabetes. Participation is always lower than 100%, biased towards participants with higher socioeconomic status and differentially so in cases and controls because of typically lower participation among controls than cases (23, 24). Severe selection and/or recall bias is therefore often present in case-control studies of socioeconomic status and type 1 diabetes. Many studies with a main aim of relating socioeconomic status to type 1 diabetes had used area-based socioeconomic status. Ecological studies are vulnerable to distinct biases that cannot be mitigated by adjustment for confounding (25). The larger and more heterogeneous the geographical area on which an individual’s socioeconomic status is attributed, the larger the potential for very strong biases that may even reverse the direction of associations or causal effects existing at the individual level.

The general lack of adjustment for ethnicity and immigration status in most studies also represent an important problem when attempting to interpret the literature (2). We excluded studies with prevalent type 1 diabetes, as type 1 diabetes in a child may influence parental socioeconomic status (26).

### The wider context of social inequality in child health

Many health aspects are well known to be associated with low socioeconomic status, including child mortality (27), and our review does not in any way negate this this. However, we should also not take for granted that all aspects of health are caused by or predicted by low socioeconomic status. Social inequality in child health represents separate methodological challenges, and it is important to differentiate between studies of objective health outcomes that are not likely to be influenced by parents’ reports or behaviour that may influence the likelihood of their child receiving a diagnosis which may create bias in studies of child health. A previous review of childhood cancer risk documented methodological weaknesses and inconsistencies in the literature similar to what we have documented here for type 1 diabetes (28). It is possible that aspects of socioeconomic status have context dependent effects. A recent study of city dwellers in high-income European countries reported higher circulating levels of several toxicants in children and their mothers with higher socioeconomic status, perhaps contrary to expectation (29).

Childhood type 1 diabetes is a well-defined disease for which underdiagnosis is not a likely problem, at least in middle- and high-income settings. It is well documented that low socioeconomic status is associated with poorer glycaemic control and comorbidities in patients with type 1 diabetes (30, 31). However, the latter is an entirely different research question that what we have addressed in the current review.

### Practical implications for future studies

Given the relatively weak and inconsistent associations between socioeconomic status and risk of type 1 diabetes documented here and the many layers of methodological problems discussed above, additional ecological studies are not likely to advance the field. Future studies should aim for prospective designs, possibly registry-based studies with complete population coverage. Furthermore, large sample size is important for sufficient power to detect the likely weak to moderate strength of associations, or conclusively rule out associations. We further recommend to avoid categorizing indicators of socioeconomic status too coarsely, and to allow for potential non-linear associations in the analysis. Finally, use clearly defined individual level socioeconomic status indicators (area-based indicators could be used together with individual level indicators in multilevel analyses).

### Strengths and imitations of the review

We have comprehensively reviewed a large literature that was scattered and sometimes difficult to identify because socioeconomic status was not necessarily part of the main aim of the study. Most studies were from middle- or high-income countries. We limited our review to childhood-onset type 1 diabetes to make interpretation of parental socioeconomic status most relevant. Type 1 diabetes may occur at any age, and socioeconomic status may have different effects in young adults. While a few studies have also included young adults (e.g. Bruno (22)), the person’s own indicator of socioeconomic status may become increasingly relevant with increasing age.

### Conclusion

We conclude that there is a need for more high-quality studies and that the existing literature does not suggest a major and consistent role of socioeconomic status as a risk factor for type 1 diabetes.

## Data Availability

All articles included in the manuscript are cited. A list of all papers included in the manuscript could be sent after request to corresponding author.

## Acknowledgments

We thank the following colleagues for commenting on an earlier draft of this manuscript: Siri E. Håberg, NIPH; Inger J. Bakken, Directorate of Health, Norway; Torild Skrivarhaug, Oslo University Hospital; Geir Joner, Oslo University Hospital.

## Abbreviations

CI: confidence interval

