## Supplemental information for "Is socioeconomic status associated with risk of childhood type 1 diabetes? A comprehensive review"

### Methods

#### *Search terms*

We searched PubMed using the following search terms:
(type 1 diabetes [Title] OR Insulin Dependent Diabetes [Title] OR Insulin-Dependent [Title] OR IDDM[Title] OR childhood diabetes [Title] OR juvenile onset diabetes[Title])
AND
(incidence [Title/Abstract] OR incident [Title/Abstract] OR new cases [Title/Abstract])
AND
(social class [Title/Abstract] OR socioeconomic [Title/Abstract] OR socio-economic [Title/Abstract] OR socio-demographic [Title/Abstract] OR social [Title/Abstract] OR education [Title/Abstract] OR maternal education* OR parental education* OR occupation [Title/Abstract] OR deprivation [Title/Abstract])

#### *Study designs and exclusions*

Most of the ecological studies had individual level data for type 1 diabetes and possibly covariates and are thus formally semi-ecological. Fully ecological studies with gross comparison of country level socioeconomic status variables and type 1 diabetes incidence were not included. We also excluded one ecological study of time trends in type 1 diabetes incidence and country level socioeconomic status such as Gross Domestic Product within a single country (1).

We excluded studies of own specific occupation in relation to development of adult onset type 1 diabetes (2), and studies that used parental occupations as indicators of social contact, without showing data for either specific occupations or occupation-based socio-economic status (3, 4).

### Results

#### *Methodological quality and overlapping publications*

Of the 69 publications with relevant data that were reviewed in detail (Main Figure 2), we excluded from the primary review publications with design weaknesses or if multiple publications represented at least partially the same observations. In many studies, different authors had used the same or highly related original data but chosen different definitions of socioeconomic status or methods of analysis (excluded publications with reason listed in Table S1).

Studies using area-based indices of socioeconomic status are prone to biases depending on the size and number of areas included. The size of the area for which the average socioeconomic status indicators are assigned to each type 1 diabetes case in the publications varied from very small "neighbourhoods" with 250-1000 residents, up to very large areas such as municipality, region, or county often with 10-100 000 residents. The size and number of areas were often not clearly reported. Variation in individual level socioeconomic status, ethnicity, etc within and between such small areas may vary dramatically between areas in the same country and between countries. This variation makes comparison across studies difficult. Most publications reported only results for a composite socioeconomic status index, while others also tested several different components of such indexes for association with type 1 diabetes. Ecological studies using composite variables of area-based socioeconomic status categorized the variables in 2-6 levels (often divided in equally sized groups), while some used components such as percent unemployed in the residential area as a continuous variable.

#### *Socioeconomic status as the main aim of the publications*

It was not always clear whether socioeconomic status was a part of an *a priori* aim or only used for statistical adjustment. However, for most publications with individual level socioeconomic status, this was not part of a main aim (4 of 8 primary cohort studies and 10 of 13 case-control studies). In contrast, investigating social inequality was the main aim in most of the ecological studies (11 of 14 ecological studies, according to our judgement).

#### *Adjustment for ethnicity*

In many countries, ethnicity, immigration, or country background represent an important confounder of the socioeconomic status - type 1 diabetes association. For instance, in the Nordic countries, ethnic minorities tend to be immigrants from Asia, Africa or South America, with lower incidence of type 1 diabetes. Some of these have low incidence of type 1 diabetes and tend to have lower socioeconomic status on average.

Only three of the eight cohort studies had accounted for immigration/ethnicity by restriction or adjustment, and one of the 14 primary case-control studies had done so (although another three were done in populations where ethnic minorities represent less than 5% of the population and ethnicity was not likely a major confounder).

Eight of 14 ecological studies had not accounted for ethnicity. Six of the primary ecological studies attempted to do so by stratifying or restricting analyses by ethnicity (Ball 2014, Puett 2012, Harron 2011, Lipton 1999, Siemiatycki 1988, and LaPorte 1981, listed in Main Table 3). In some cases, e.g. Siemiatycki 1988, ethnicity could not be satisfactorily adjusted for because of different sources of ethnicity in the type 1 diabetes group and the general population.

#### *Socioeconomic status indicators used in the publications*

Parental education and parental occupation were the most studied socioeconomic status variables in publications with individual level data. Most publications used three categories for maternal or paternal education, while some categorised children's socioeconomic status based on the highest level of education of either the mother or the father (indicated in our results tables with "parental education"). Reporting parental education does not allow disentangling of potential difference between maternal and paternal education socioeconomic status with respect to their association with risk of type 1 diabetes. Furthermore, it was not always clear whether education referred to the highest level initiated or completed.

Many studies used parental occupation as an indicator of socioeconomic status, mostly paternal occupation. While this was very frequently used in older literature, it is becoming increasingly difficult to clearly categorise many occupations (5). The number of categories of social status based on parental occupation in the publications ranged from two to seven.

#### *Details of studies relating farming to childhood type 1 diabetes*

we found three studies of some relevance for parental farming. The first was a questionnaire-based comparison of 242 prevalent cases with type 1 diabetes and hospital-based controls in Germany where the authors reported no significant association between living on a farm or regular contact with farm animals, and risk of type 1 diabetes (6). The second was a Finnish questionnaire-based cohort study of 5805 children of which 35 reported type 1 diabetes diagnosed before 18 years of age. Rural living on different types of farms were not association with type 1 diabetes, but there were too few children with type 1 diabetes for this study to be informative (7). The third was a Finnish cohort of children with high-risk type 1 diabetes genotypes of whom 51 developed type 1 diabetes reported family occupation in farming (odds ratio:1.58, 95% CI 0.36-6.96) or child contact with farm animals in the first year of life (odds ratio:0.79, 95% CI 0.12-5.15) not to be significantly associated with risk of type 1 diabetes (8).

#### *Details on case-control study on parental occupational exposure*

We found only publication investigating parental occupational exposure (9). This was a case-control study in Belgrade from which it was difficult to draw firm conclusions. The authors reported that "Parents of diabetic children were significantly more frequently occupationally exposed to radiation, petroleum, and its derivates, organic solvents, dyes and lacquers" (9). These authors also reported the same data as part of a multivariable analysis in Sipetic 2005 (10), now stating that "mother's and father's specific occupational exposure to radiation, low temperature, petroleum and its derivates, metallic dust, asbestos, steel, organic solvents, dyes and lacquers" were more frequent in children with diabetes compared to controls (controls were children hospitalised with skin diseases). These publications describe the same case-control study from Belgrade as Sipetic 2004 presented in Table 2). Sipetic 2004 reported five categories of maternal and paternal occupations, with no overall significant association with type 1 diabetes. Parents who were farmers were too few for the study to be informative (zero cases and 1% of controls). Approximately 9% and 7% of mothers of cases and controls, respectively, were miners/industrial workers. 10 vs 8% of fathers were miners or industrial workers among 105 cases and 210 controls (11).

Table S1.  Characteristics of 34 publications not considered in the primary review*

| First author, year published (reference) | Reason for excluding from main review* | Period T1D diagnosed | No. w/T1D (age)† | Place | SES variables ‡ | Design§ | Direction of association‖ | Result, comment |
|---|---|---|---|---|---|---|---|---|
| **Cohort studies (10 publications)** | | | | | | | | |
| Antvorskov 2020 (12) | Overlap Clausen 2016 (main Table 1) | 1997-2014 | 322 | Denmark | Parental educ./occup. | Cohort | (-) | High educ/occup. lower T1D risk(p=0.08) |
| Metsälä 2018 (13) | Many missing and mix of educ. and occup. | 1981-2009 | 9541 (<16y) | Finland | Maternal occup. (or education if occup. missing) | Case-cohort | No association | No association (uRR=0.99-1.00 for upper 2 vs lower). Missing: RR=2 |
| Virk 2016 (12) | Overlap Clausen 2016 (main Table 1), age 5-35 | 1980-2010 | 5732 (5-31y) | Denmark | Maternal education | Cohort | – | Higher mat. educ. lower T1D risk |
| Hussen 2015 (14) | Overlap Khashan 2015 (main Table 1) | 1992-2009 | 5771 (<18y) | Sweden | Parental‡ education | Cohort | Non-linear | Parental educ middle cat: aIRR=1.15 vs 1.0 in high and low if Nordic parents |
| Hussen 2013 (15) | Overlap Khashan 2015 (main Table 1) | 1969-2008 | 17717 | Sweden | Parental‡ education | Cohort | (+) | Higher parent educ. slightly higher T1D risk, but inconsistent in subgroups |
| Nygren 2015 (16) | <100 T1D cases | 1997-2012 | 58 | Sweden (south east) | Mat. and pat. educ. | Cohort | ? | Complex pattern by combination of maternal and paternal education |
| Hjern 2008 (17) | Overlap Khashan 2015 (main Table 1) | 1987-2003 | 3225 | Sweden | Mat. educ., head of household SES (occupation), other | Cohort | + | Higher maternal education higher T1D risk (non-linear). |
| Waldhoer 2008 (18) | Data not reported in detail ("not significant") | 1989-2005 | 444 (<5y) | Austria | Maternal education (and marital status) | Cohort | ? (not significant) | SES variables categories not clearly stated |
| Hyppönen 2001 (19) | <100 T1D cases | 1966-1997 | 80 (<31y) | Finland (Oulu and Lappland) | Maternal education | Cohort | – | More than basic vs none or basic: uRR=0.51, 95%CI: 0.35-1.00 |
| Jones 1998 (20) | Overlap Ievins 2007 (main Table 2) | 1965-1987 | 315 | UK (Oxfordshire and West Berkshire) | SES based on parental occupation | Cohort | No clear association | No association |

| | | | | | | | | |
|---|---|---|---|---|---|---|---|---|
| **Case-control studies (4 publications)** | | | | | | | | |
| Baruah 2011 (21) | Single hospital, few T1D | 2000-2001 | 43 (<18y) | India (New Delhi) | Parental educ., "SES" | Case-control | No clear association | No association |
| Marshall 2004 (22) | Prevalent T1D | 1982-1998 | 196 (<16y) | UK (Lancashire and Cumbria) | Paternal academic qualifications (yes/no) | Case-control | + | aRR=2.0, 95% CI (1.10-3.60) |
| McKinney 2001 (23) | Overlap McKinney 1997 (main Table 2) | 1993-1994 | 220 | UK (Yorkshire) | Mat. educ., area composite index (Townsend) at birth | Case-control | – | Higher maternal education lower T1D risk |
| Tai 1998 (24) | Friend controls matched for education | 1984-1993 | 117 (<30y) | Taiwan (Taipei city) | Family income (but freq matched for education) | Case-control | + | Higher family income lower T1D risk (matched for education) |
| Mayer 1988 (25) | Overlap Lawler-Heavner 1994 (main Table 2) | 1978-1985 | 268 | US (Colorado) | Maternal educ., family income | Case-control | No clear association | Maternal education no association; Higher family income lower T1D risk |
| **Other design with individual level SES (3 publications)** | | | | | | | | |
| Metcalfe 1992 (26) | Different source of SES for cases and controls (overlap Wadsworth 1997; main Table 2) | 1988 | 1175 | UK + Ireland | Parental occupation social class | T1D vs national reference data | + | High SES paternal occupation higher T1D risk |
| Tarn 1983 (27) | Prevalent T1D, limited details (Letter), census not directly comparable | 1982 | 186 (<26y) | UK (near London) | Social class (based on occupation) | Hospital records + census | + | Highest social class higher T1D risk, but not consistent (non-linear) |
| Debono 1983 (28) | Prevalent T1D, limited details (Letter), census not directly comparable | 1981 | 62 | UK (Southampton) | Social class (based on occupation) | Hospital records + census | + | Highest social class higher T1D risk, threshold association |
| **Ecological studies§ (16 publications)** | | | | | | | | |
| Rafferty 2021 (29)** | Results only for age range 0-60 year | 2008-2018 | <7857 (0-60y) | Wales | Composite area index | Ecological | - | Higher SES lower T1D risk |
| Grigsby-Toussaint 2010 (30) | Unclear SES indicator, overlap Lipton 1999 (main Table 3) | 1994-2003 | 765 (<17y) | US (Chicago) | Neighbourhood income "diversity" (past 3 decades) | Ecological | ? | Higher risk in emerging low-income neighbourhoods, but not consistent |

| | | | | | | | | |
|---|---|---|---|---|---|---|---|---|
| Torres-Aviles 2010 (31) | SES and T1D shown only for 13 of 52 districts | 2000-2005 | 603 | Chile (Santiago) | Area composite index | Ecological | (+) | Higher SES areas higher T1D risk (unclear statistical significance) |
| du Prel 2007 (32) | Overlap Castillo-Reinado 2020 (main Table 3) | 1996-2000 | 2499 | Germany (North Rhine-Westphalia) | Area income, education, etc and composite index | Ecological | – | Higher SES index lower T1D risk. No association for other area SES measures |
| Haynes 2007 (33) | Overlap Ball 2014 (Main Table 3) | 1984-1998 | 588 | Western Australia | | Ecological | No association | No association (analysis restricted to European origin) |
| Haynes 2006 (34) | Overlap Ball 2014 (main Table 3) | 1985-2002 | 1143 | Western Australia | Area composite index | Ecological | + | Higher SES area index higher T1D risk |
| Feltbower 2005 (35) | Overlap Harron 2011 (main Table 3) | 1986-1998 | 1551 | UK (Yorkshire) | Area composite index (Townsend) | Ecological | + (non-linear) | Higher SES area slightly higher T1D risk |
| Evans 2000 (36) | Prevalent T1D | 1993 | 792 | Scotland (Tayside) | Area composite index (Carstairs) | Ecological | No clear association | Unclear association |
| Baumer 1998 (37) | Prevalent T1D | 1994 | 801 | UK | Area composite index (Townsend) | Ecological | No clear association | Unclear association |
| Staines 1997 (38) | Overlap Harron 2011 (main Table 3) | 1978-1990 | 1490 (<17y) | UK (Yorkshire) | Area composite index (Townsend) | Ecological | (+) | Less deprived slightly higher T1D risk, but n.s. after adjustment |
| Patterson 1996 (39) | Overlap Cardwell 2006 (main Table 3) | 1989-1994 | 462 | Northern Ireland | Area composite index (and components) | Ecological | + | High SES area slightly higher T1D risk |
| Tuchinda 1992 (40) | < 100 T1D cases | 1985 | 35 | Thailand | Family education and SES (unclear definition) | Ecological/Unclear | (–)? | Unclear ascertainment of cases |
| Patterson 1992 (41) | Overlap Patterson 1991 (main Table 3) | 1977-1983 | 2125 (<19y) | Scotland | Area composite index (Carstairs) | Ecological | + | Higher SES area higher T1D risk, but assoc. varied by urban/rural |
| Colle 1981 (42) | Overlap Siemiatycki, 1988 (Main Table 3) | 1971-1978 | 588 (<17y) | Canada (Montreal) | Area average family income at diagnosis | Ecological | + | High family income higher T1D risk |
| West 1979 (43) | Overlap Siemiatycki, 1988 (Main Table 3) | 1971-1977 | 522 (<17y) | Canada (Montreal) | Small residential area based (average family income) at diagnosis | Ecological | + (non-linear) | Higher average area income higher T1D risk (non-linear) |
| Christau 1977 (44) | Study area divided geographically (two areas) | 1970-74 | 474 | Denmark, Copenhagen | North (wealthy) vs South | Ecological | - | Higher SES lower T1D risk |

u/aRR: Unadjusted (u) or adjusted (a) relative risk (or odds ratio, incidence rate ratio or hazard ratio) comparing higher versus lower socioeconomic status.
* Studies here had relevant data on SES and type 1 diabetes but were not included among the primary studies (reported in main Tables 1-3). Reasons are given in the second column of this table. The review strategy was to include in the primary review only the most recent and/or most informative in case (partially) overlapping data were reported in several publications (marked with “overlap author year”, where this refers to the primary publications. Some publications were in the present table because of methodological weaknesses or including age groups other than 0-15 years. Sorted by design, and then by publication year.
† Age (years, y) at diagnosis of type 1 diabetes (T1D) less than 15 years unless otherwise specified. Cases with available data on socioeconomic status are reported.
‡ Socioeconomic (SES) variables. Parental educ (education) or occupation refers to highest level within the pair of maternal or paternal education, as opposed to separate data for mat. (maternal) or pat. (paternal) educ.
§ Study designs: There are many variants of both cohort, case-control and (semi-) ecological designs. Case-control or case-cohort designs based on registries were regarded of equal quality as cohorts. Ecological design here refers to semi-ecological designs where type 1 diabetes (and some other variables) are available at the individual level, but socio-economic status (SES) variables are only available as an average at the place of residence at the time of diagnosis of type 1 diabetes (T1D). The size of the area for which the average SES indicators are assigned to each T1D case may vary across studies from very small “neighbourhoods” with 250-1000 residents, up to very large (low resolution) such as municipality, region or county often with 10-100 000 residents. Variation in individual level SES, ethnicity etc within and between such small areas may vary dramatically between areas in the same country and between countries. This variation makes comparison across studies very difficult. (“full” ecological designs, with gross comparison of SES indicators and T1D incidence across countries were not included here).
|| +: increased socioeconomic status (SES) was associated with higher risk of type 1 diabetes. –: Increased SES was associated with lower risk of type 1 diabetes. Lower degree of deprivation and unemployment means higher SES. Socioeconomic status based on occupation typically used in the UK are often labelled with roman number I for high social class and III, IV or V refers to lower SES.
** References listed in Supplemental reference list below
